# Quantitative imaging of the central lymphatic system with spectral CT iodine mapping: a feasibility study in swine

**DOI:** 10.64898/2026.05.06.26352364

**Authors:** Leening P. Liu, Alexey Gurevich, George McClung, Maxim Itkin, Peter B. Noël

## Abstract

**Purpose:** Imaging of the central lymphatic system enables characterization of patient-specific lymphatic anatomy and accurate localization of leaks. Advancements in CT technology, particularly spectral CT, can enhance CT lymphangiography (CTL) with improved visualization and quantification. This study aimed to assess the feasibility of spectral CTL in both static and dynamic scans.

**Materials and Methods:** 50% diluted iodinated contrast was injected into the bilateral superficial inguinal lymph nodes of a pig. The pig was scanned with a dual-layer spectral CT every 60 seconds for 10 minutes. To optimize contrast and visualize peristalsis, a second animal was injected with 25% and 10% diluted contrast and scanned dynamically 4 and 6.25 minutes after contrast injection. Conventional images and iodine maps were reconstructed to calculate the contrast-to-noise ratio (CNR). Additionally, the iodine density was measured adjacent to the lymphovenous junction to show fluctuations from peristalsis and contrast washout.

**Results:** Iodine maps, compared to conventional images, separated the contrast-filled central lymphatic system from surrounding soft tissue and increased CNR to 895 compared to 43 with conventional images. 25% diluted contrast provided the best balance between visualization and quantification of the central lymphatic system, showing high and low iodine density regions corresponding to peristalsis. Iodine density peaked at 15.4 ± 0.6 mg/mL and decreased to 2.0 ± 0.1 mg/mL at 10.5 minutes.

**Conclusion:** Spectral CTL not only improves visualization of the central lymphatic system compared to CTL but also provides quantitative information for physiological characterization of lymphatic disease that can enhance current subjective assessment.

**Research highlights:**

- Iodine maps from spectral CT lymphangiography separated contrast-filled lymphatic structures from surrounding soft tissue and provided better contrast-to-noise compared to conventional images.
- Spectral CT lymphangiography enabled quantification of contrast in the central lymphatic system that demonstrated contrast washout and may be utilized for physiological characterization of disease.
- Dynamic spectral CT imaging of the lymphatic system visually showed peristalsis in the thoracic duct and was further reflected in quantitative iodine density measurements.

## Introduction

The central lymphatic system is a vital component of the lymphatic system that is responsible for maintaining fluid homeostasis by returning interstitial fluid and proteins to venous circulation. Disruption of lymphatic integrity due to surgical trauma or congenital lymphatic disorders can result in lymphatic leaks. These leaks may manifest as severe clinical conditions, including chylothorax, chylous ascites, and protein⍰losing enteropathy (1–4). Lymphatic leaks can result in profound malnutrition, dehydration, electrolyte imbalances, and immune cell loss (1,4). As a result, detection of leaks of the thoracic duct and its variants is central to both diagnosis and management. Characterization of patient-specific lymphatic anatomy and accurate localization of the leak can determine the success of non-surgical management, such as thoracic duct embolization (TDE) (5–7). Despite the increasing clinical volume of lymphatic interventions, comprehensive, reproducible, and quantitative imaging of the thoracic duct and central lymphatic vessels remains technically challenging (8).

Imaging of the central lymphatic system has ranged from intranodal lymphangiography to dynamic contrast-enhanced magnetic resonance lymphangiography (DCMRL) (9). DCMRL has become increasingly popular as a preprocedural planning tool because of its three-dimensional acquisition, good tissue resolution, and use of water-soluble contrast, which is significantly less viscous than oil-based contrast utilized in intranodal lymphangiography (9,10). However, logistical and technical challenges hinder its widespread use: MRI has limited global availability compared to CT, is expensive, requires long scan times, and is relatively contraindicated in patients with paramagnetic implants or claustrophobia. Furthermore, ultrasound-guided needle placement for DCMRL must take place outside of the MRI suite, increasing the risk of needle dislodgement during patient transport (10,11).

In comparison to DCMRL, CT lymphangiography (CTL) with water-soluble iodinated contrast offers more accessible and faster imaging of the central lymphatic system (8,12). Though CTL requires radiation exposure, design of the dose-optimized protocols, such as stepwise propagation scanning, have enabled a reduction in radiation exposure while maintaining image quality (12). However, even with optimized CTL, lymphatic opacification can be heterogenous and depend on time. Additionally, diagnostic interpretation of the central lymphatic system on conventional reconstructions is complicated by the complex anatomical background and remains only qualitative. Development of objective quantitative metrics may facilitate more accurate diagnosis of branched lymphatic vessel ruptures and enhance understanding of flow dynamics (12,13).

Spectral CT enables improved contrast visualization and quantification ideal for imaging the central lymphatic system. Even though a few studies have implemented spectral CT to evaluate the distribution of contrast and extravasation after lymphangiography (14), they did not leverage the benefits of spectral CT, particularly iodine maps. Spectral CT uses attenuation measurements from high⍰ and low⍰energy spectra to perform basis material decomposition and reconstruct iodine maps. These iodine maps show iodinated contrast within structures, eliminating all of the anatomical background. In addition, quantitative values on iodine maps represent the concentration of iodine, which enables both quantitative measurement and evaluation of physiological processes, such as vascularity. For lymphatic imaging, complete separation of iodine signal from the background soft tissue, bone, and surgical material improves visualization and can be advantageous for intranodal CTL where detection of low contrast concentration structures and flow is critical to accurate diagnosis (11). Quantification of the contrast within the thoracic duct is also feasible with iodine maps. Compared to conventional Hounsfield Units (HU), iodine maps are quantitatively reliable, largely agnostic to key protocol parameters, such as tube voltage (15–18). As a result, quantitative metrics derived from iodine maps may enable characterization of both washout (19–21) and flow dynamics (13) of the central lymphatic system. Thus, the aim of this study is to evaluate the feasibility of spectral CTL for visualizing and quantifying contrast in the central lymphatic system. The feasibility was assessed at different contrast levels and with both static and dynamic imaging.

## Methods

### Animal procedures

All procedures were reviewed and approved by the University of Pennsylvania’s Institutional Animal Care and Use Committee (protocol #807306). Two four-month old male Yorkshire pigs (50 and 54 kg) were utilized: one to evaluate overall feasibility of spectral CTL and one to optimize contrast parameters and assess feasibility of dynamic spectral CTL. Both animals were sedated with an intramuscular injection of butorphanol (Torbugesic, Zoetis, Parsippany, NJ, 0.1-0.5mg/kg) and dexmedetomidine (Dexdomitor, Zoetis, Parsippany, NJ, 0.01-0.05 mg/kg). After sedation, an auricular catheter was placed for intravenous access. Anesthesia was induced with intravenous ketamine (Ketaset, Zoetis, Parsippany, NJ, 1-5 mg/kg) and/or propofol (PropoFlow, Zoetis, Parsippany, NJ, 1-10 mg/kg). The animals were then placed in a dual-layer spectral CT (Spectral CT 7500, Philips Healthcare, Amsterdam, Netherlands). The animals were then intubated and maintained on a mixture of isoflurane (IsoFlo, Zoetis, Parsippany, NJ, 1-5%) in air during the procedure. In both animals, the bilateral superficial inguinal lymph nodes were accessed with “Hook” needles (SureAx-Hook, SureAx Medical, San Diego, CA) for contrast injection.

### Data acquisition

To investigate the overall feasibility of spectral CTL, a pre-contrast, full body static scan of animal A was performed at a tube voltage of 140 kVp and an exposure of 300 mAs (Figure 1A). Additional acquisition parameters can be found in Table 1. Iodinated contrast (Isovue-300, Bracco Diagnostics, Princeton, NJ) was diluted 1:1 with saline (0.9% Sodium Chloride Irrigation USP, Baxter, Deerfield, IL) to create a 50% iodinated contrast solution with an iodine concentration of approximately 150 mg/mL. Diluted iodinated contrast was then injected with a power injector (MEDRAD Stellant FLEX, Bayer AG, Leverkusen, Germany) at a volumetric rate of 0.1 mL/s and a dose of 0.4 mL/kg to match our institution’s injection protocol for MR lymphangiography. Scans were then performed every 70 seconds for 10.5 minutes to visualize the central lymphatic system over time (Figure 1A).

**Figure 1.**
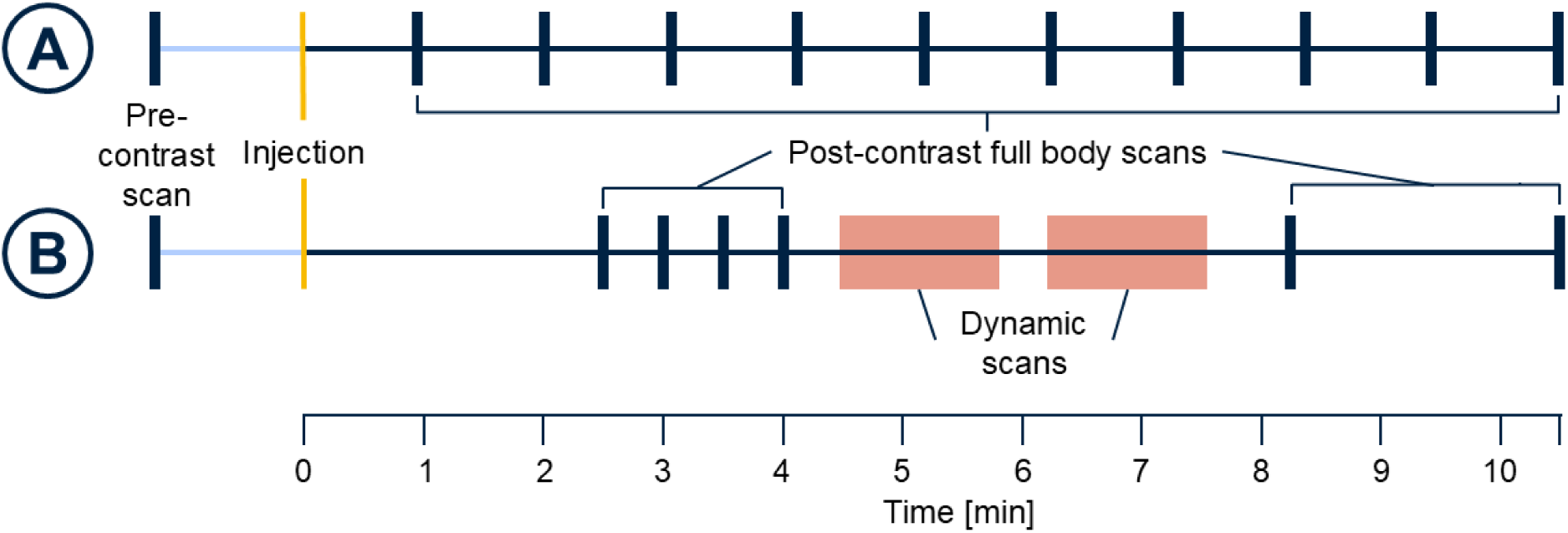
Timeline for spectral CTL. To evaluate the overall feasibility of spectral CTL, the first animal was imaged before contrast injection and after contrast injection approximately every 70 seconds for 10.5 minutes (A). To optimize contrast protocol and assess imaging of peristalsis, the second animal was imaged before contrast injection followed by post-contrast full body scans and dynamic scans to capture the peristalsis of the central lymphatic system (B). This procedure was performed with both 10% and 25% diluted contrast.

**Table 1.** Acquisition and reconstruction parameters.

|  | **Animal A** | **Animal B** |  |
| --- | --- | --- | --- |
|  | **static** | **static** | **dynamic** |
| **Tube voltage [kVp]** | 140 | 140 | 120 |
| **Collimation** | 128 x 0.625 | 128 x 0.625 | 128 x 0.625 |
| **Exposure [mAs]** | 300 | 250 | 200 |
| **CTDIvol [mGy]** | 34 | 28 | 30 |
| **Scan type** | helical | helical | axial |
| **Pitch** | 0.61 | 0.61 | - |
| **Rotation time [s]** | 0.5 | 0.272 | 0.272 |
| **Denoising level (iDose<sup>4</sup>)** | 4 | 4 | 4 |
| **Reconstruction kernel** | B | B | UB |
| **Reconstruction diameter [mm]** | 350 | 350 | 350 |
| **Slice thickness [mm]** | 0.67 | 0.8 | 0.8 |
| **Matrix size** | 512 | 512 | 512 |
| **Pixel spacing, x and y [mm]** | 0.68 | 0.68 | 0.68 |

To evaluate the feasibility of dynamic spectral CTL, animal B was scanned at a tube voltage of 140 kVp and an exposure of 250 mAs prior to contrast injection (Figure 1B). A faster rotation time of 0.272 seconds was utilized to better match the minimum rotation time for dynamic scans. In comparison to the first animal, 25% and 10% iodinated contrast solutions were created by diluting iodinated contrast with saline. For each contrast solution, diluted iodinated contrast was injected with a power injector at a volumetric rate of 0.1 mL/s and dose of 0.2 mL/kg to reduce contrast load and reflect the contrast protocol in Ahn et al (12). Two and a half minutes after contrast injection, the animal was scanned every 30 seconds for 1.5 minutes to ensure opacification of the central lymphatic system and contrast washout (Figure 1B). With contrast washing out, sets of dynamic scans of the upper thorax adapted from a brain perfusion protocol were performed at 4.5 and 6.25 minutes. The animal was scanned at a tube voltage of 120 kVp and an exposure of 200 mAs every 5 seconds for 16 scans, totaling 80 seconds (Table 1, Figure 1B). The mild reduction in exposure compared to static scans was implemented to capture additional scans in each dynamic scan set. At 8.25 and 10.5 minutes, additional full-body scans were performed to visualize washout of the contrast (Figure 1B).

### Image analysis

For all scans, conventional images and iodine maps were reconstructed with a field of view of 350 mm. All other reconstruction parameters can be found in Table 1. To fully visualize the central lymphatic system, coronal maximum intensity projections (MIP) through 50 mm were generated for both conventional and iodine density maps. In axial images across time points, a circular region of interest (ROI) with a diameter of 4 mm was placed at the lymphovenous junction. The mean and standard deviation (noise) in ROIs were measured in both conventional images and iodine maps to characterize washout. Additional ROIs were placed in the artifact-free soft tissue adjacent to the lymphovenous junction for each time point to calculate the contrast to noise ratio (CNR). A scatter plot was then utilized to detail the changes in iodine density during washout. To demonstrate peristalsis, the difference in iodine density between each dynamic scan (every five seconds) was calculated. For each dynamic scan series, the temporal change in iodine density was calculated at five⍰second intervals. The mean change and standard deviation were used to describe time⍰dependent variability in measured iodine concentration.

## Results

### Overall feasibility

Compared to conventional images (Figure 2A), iodine maps isolated the central lymphatic system from the background soft tissue (Figure 2BC). Pre-contrast iodine map MIPs showed residual bone and devices including a nasogastric tube for removing air in the stomach and “Hook” needles in the bilateral superficial inguinal lymph nodes (Figure 2D). After injection of 50% diluted iodinated contrast, iodine maps illustrated the full pathway from the superficial inguinal lymph nodes through the cisterna chyli and thoracic duct before finally draining into the venous system at the lymphovenous junction (Figure 2E). Iodinated contrast was first detected at 38 mg/mL, the system’s upper limit for iodine quantification. This iodine density corresponded to a CNR of 895 compared to a CNR of 43 observed in conventional images, reflecting a 20-fold increase with iodine maps. Over time, washout of iodine was present on iodine maps, which was particularly noticeable at the lymphovenous junction (Figure 2F). At 10.5 minutes, iodine concentration decreased to 5.9 ± 0.1 mg/mL.

**Figure 2.**
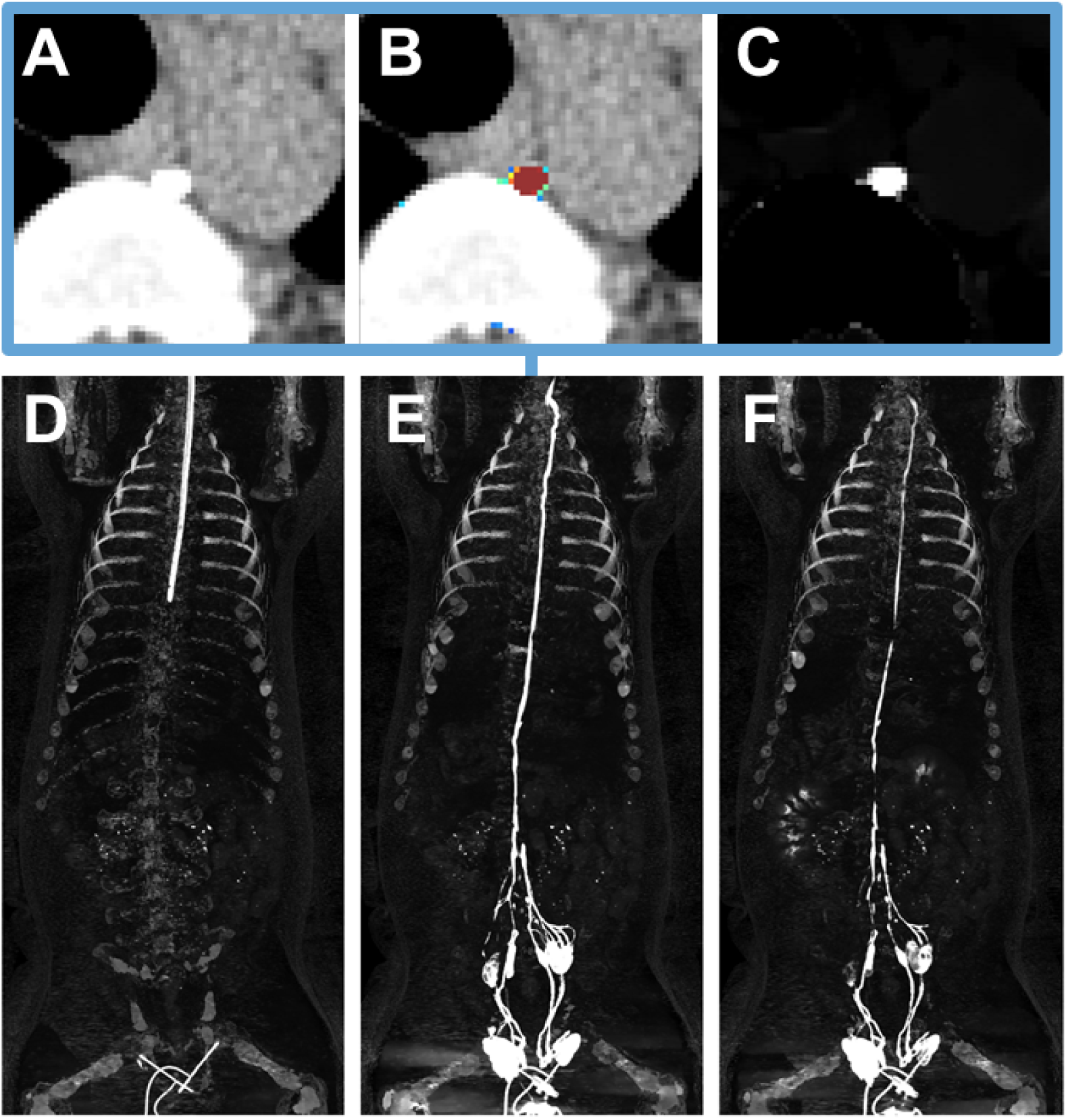
Spectral CTL with conventional images and iodine maps. Axial images demonstrate clear visibility of the thoracic duct on both conventional images (A), conventional images with iodine overlay (B), and iodine maps (C). Iodine maps showed improved contrast to noise ratio and enabled quantitative measurements. Pre-contrast maximum projection images (MIP, D) of iodine maps included initial placement of a nasogastric tube to evacuate gas from the gastrointestinal system and bilateral placement of “Hook” needles in the superficial inguinal lymph nodes. Post-contrast MIP (E) enabled visualization of the entire central lymphatic system from the superficial inguinal lymph nodes to the cisterna chyli to the lymphovenous junction. At 8.5 minutes post-contrast, MIP (F) showed washout of the contrast from the lymphatic system into the venous system. Conventional images (A) WL/WW: 40/400 HU. Iodine maps (B-E) WL/WW: 6/12 mg/mL.

### Contrast optimization

Of the different concentrations of contrast injected, 25% diluted contrast demonstrated not only adequate illumination of the central lymphatic system but also moderate iodine density values for quantification at three minutes (Figure 3). Iodine map MIPs with 25% diluted contrast (Figure 3B) showed similar opacification of the central lymphatic system compared to 50% contrast (Figure 3A). However, at 10% diluted contrast, despite the clear visualization of the distal central lymphatic system on iodine MIPs, the thoracic duct was more difficult to visualize and had an iodine density of 1.2 ± 0.1 mg/mL (Figure 3C). The resulting iodine density was not suitable for assessing flow and peristalsis and thus excluded. While the peak iodine density measured with 50% diluted contrast reached the system’s upper limit for iodine quantification, the peak iodine density at the lymphovenous junction with 25% diluted contrast measured 15.4 ± 0.6 mg/mL at three minutes post injection. The corresponding CNR measured 295 for 25% diluted contrast, which remained greater than the CNR of 43 observed in conventional images with 50% diluted contrast.

**Figure 3.**
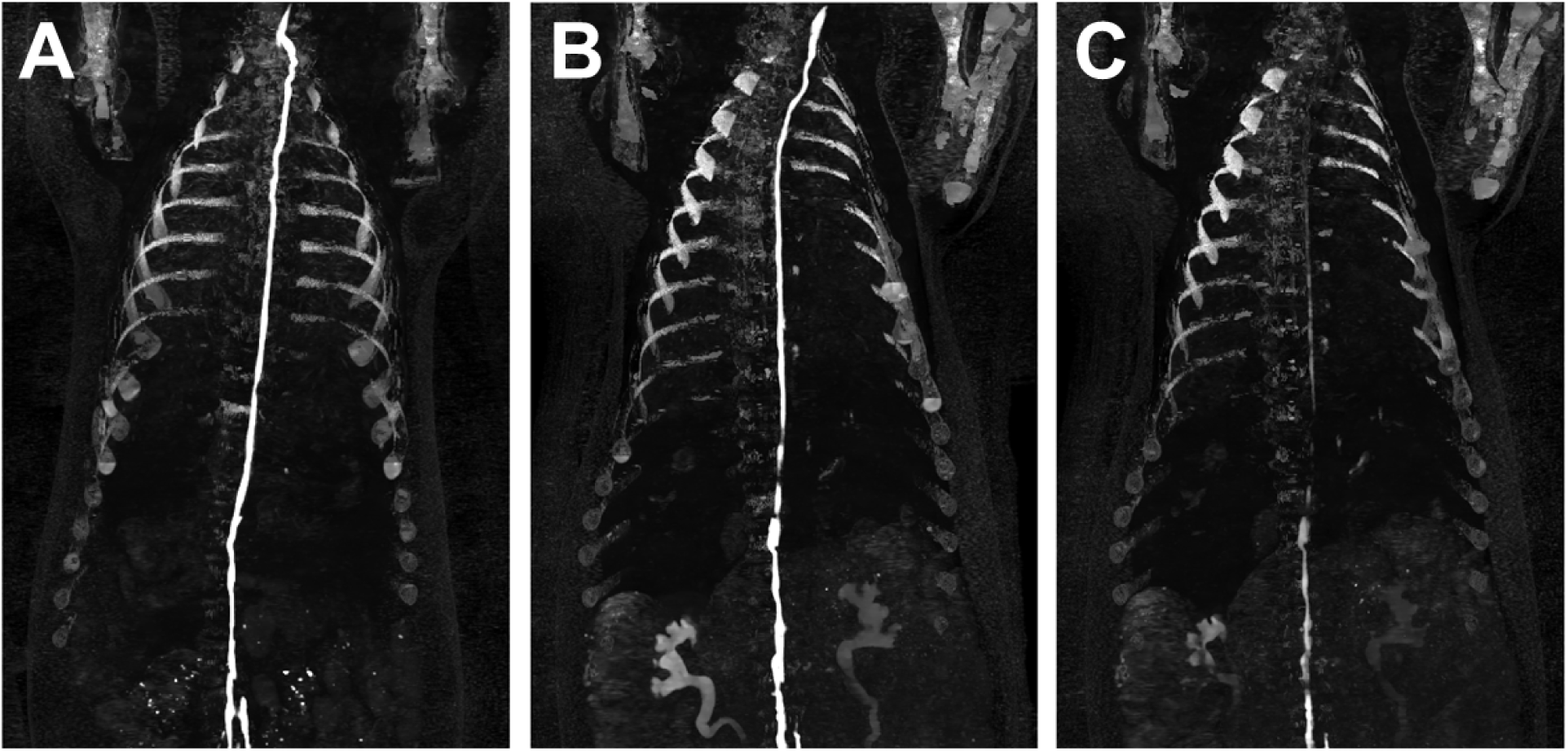
Contrast optimization for spectral CTL. Iodine map MIPs at 3 minutes obtained with 50% (A) and 25% (B) diluted contrast demonstrated full opacification of the central lymphatic system. However, with 50% diluted contrast, iodine maps exhibited iodine densities greater than the system’s upper limit of quantification, making them non-ideal for quantification. With 10% diluted contrast, MIP at 3 minutes (C) showed significant reduction in contrast and diminished visualization of the thoracic duct in the thorax. WL/WW: 6/12 mg/mL.

### Peristalsis

Dynamic images with 25% diluted contrast visually and quantitatively reflected the peristaltic nature of the thoracic duct (Figure 4). Initially, from 4:30 to 4:45, iodine densities within the thoracic duct were too high to visualize this effect at the window shown in Figure 4. As contrast washed out, iodine map MIPs showed areas of high and low iodine densities along the length of the thoracic duct distal to the lymphovenous junction. The resulting beading of contrast is first the most apparent at 4:50 where there were three areas of low iodine density, corresponding to sites of peristalsis. Five seconds later, the sections of low iodine density were no longer present but reappeared at approximately the same locations at 5:05, 5:15, 5:30, and 5:45. In the second set of dynamic images, the same effect was present but more subdued due to contrast washout. The presence of peristalsis was also demonstrated in quantitative measurements (Figure 5). During the first set of dynamic scans from 4:30 to 5:45, iodine density ranged from 9.3 ± 0.9 to 6.8 ± 0.3 mg/mL, highlighting washout. The difference in iodine density every five seconds averaged -0.06 ± 0.94 mg/mL and ranged from -2.0 to 1.8 mg/mL. Similarly, the average difference in iodine density every five seconds for the second set of dynamic scans was -0.04 ± 0.68 mg/mL, showing the peristalsis in the thoracic duct.

**Figure 4.**
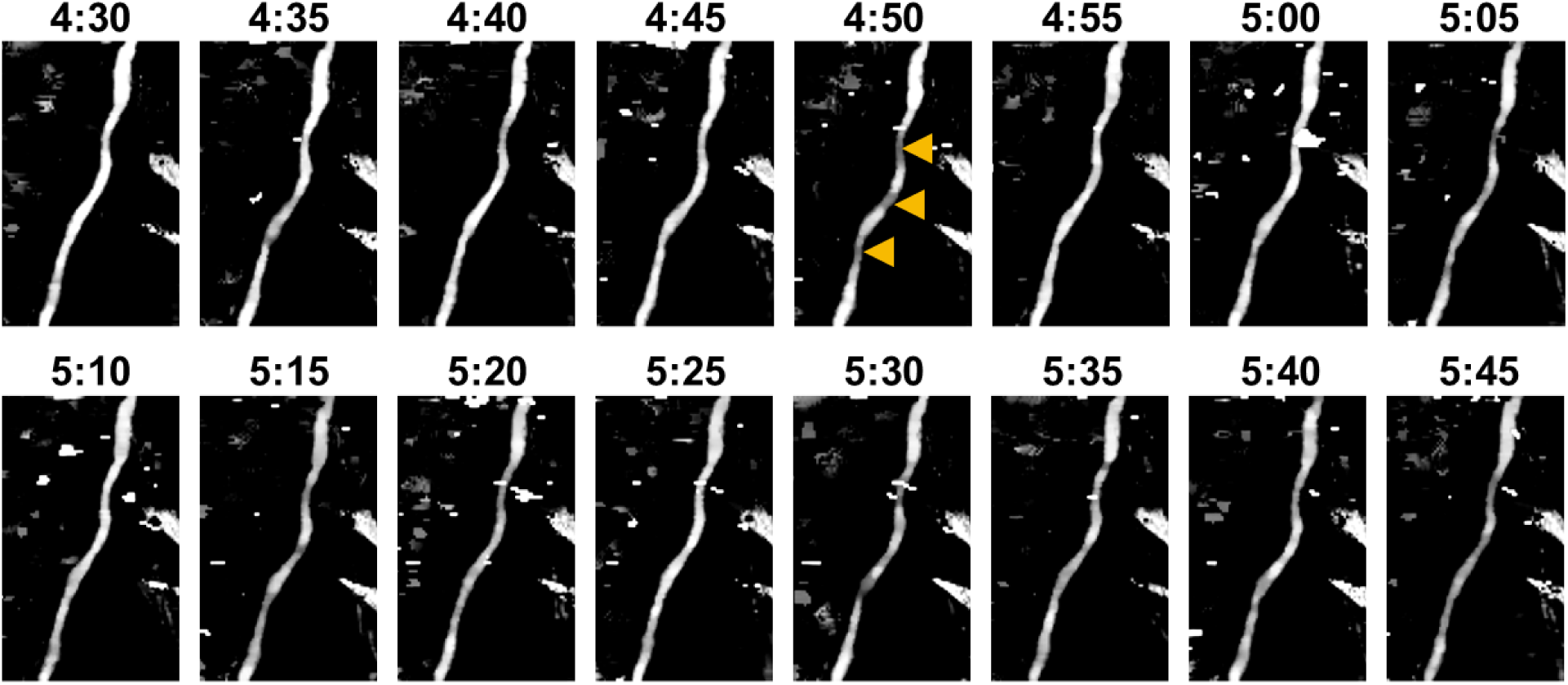
Dynamic imaging of peristalsis in the thoracic duct. Iodine map MIPs obtained at each time point exhibited high and low iodine density along the segment of the thoracic duct. Across time points, these high and low iodine density sections shifted along the thoracic duct, demonstrating the peristaltic nature of the thoracic duct. Yellow arrowheads indicate sites of peristalsis. WL/WW: 6/6 mg/mL.

**Figure 5.**
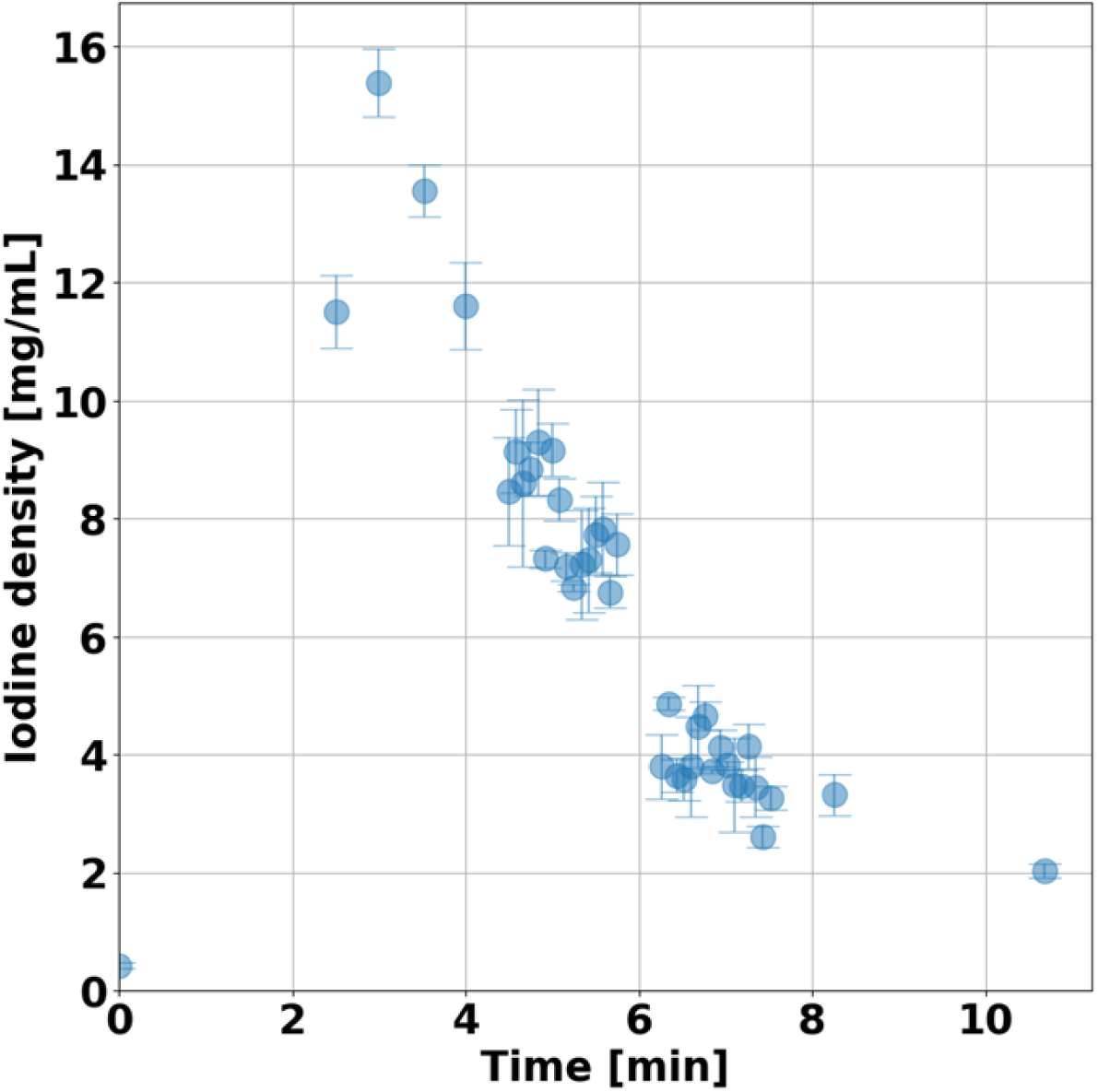
Iodine density adjacent to the lymphovenous junction over time with 25% diluted contrast. Iodine density peaked at 3.5 minutes and decreased with time as contrast washed out of the central lymphatic system into the venous system. Iodine density measured in dynamic scans (4.5 – 7.5 minutes) demonstrated fluctuations that correspond to the peristaltic nature of the thoracic duct.

## Discussion

In vivo imaging of the central lymphatic system not only demonstrated the feasibility of spectral CTL but also highlighted its benefits over conventional CTL. Iodine maps generated with spectral CT improved visualization of the central lymphatic system compared to conventional images by isolating contrast-enhanced structures and eliminating soft tissue background. Additionally, quantification of the contrast in the thoracic duct showed contrast washout that can be utilized to characterize flow and the peristalsis of the thoracic duct. Both visualization and quantification with spectral CTL can enhance non-invasive evaluation of lymphatic disorders prior to intervention.

Compared to DCMRL, spectral CTL provides maps that enable quantification of washout, flow, and peristalsis. Even though DCMRL has been critical to the diagnosis of lymphatic disorders, characterization of these disorders has been largely qualitative. Quantitative physiological⍰related metrics, such as flow, may facilitate disease stratification and guide clinical management. Over the last few years, a few studies have investigated non-invasive measurements of flow with both DCMRL (22) and CTL (13). These studies implemented automated segmentation and binary thresholding to determine changes in volume, which do not capture nuanced changes in concentration (13,22). An additional method for quantifying flow in CTL utilized changes in attenuation as a surrogate for iodine concentration and captured the entire thoracic duct as a single compartment (13). Although this study did not derive formal flow metrics, direct measurement of iodine density enabled assessment of contrast distribution and time⍰dependent changes along the thoracic duct. To ensure reliable quantification along the thoracic duct, a 25% diluted contrast is recommended instead of the 50% diluted contrast utilized in DCMRL because it resulted in values above 38 mg/mL, the system’s upper limit of quantification. As demonstrated in this study, the use of 25% diluted contrast also enabled visualization and quantification of peristalsis. In order to better capture the frequency and motion of peristalsis, an interval shorter than five seconds between perfusion scans is required. These quantitative capabilities of spectral CT align with the broader evolution towards the development and incorporation of physiological-related metrics for understanding pathophysiology and characterizing disease.

In addition to the quantitative capabilities, spectral CTL provides significant benefits for lymphatic imaging. First, spectral CTL improves conspicuity of the thoracic duct anatomy and variants critical for interventional planning. Iodine maps show only contrast-enhanced structures, removing the complex anatomical background and confirming the presence of low-contrast pathological findings, such as leaks and abnormal collaterals. Second, spectral CTL provides a higher spatial resolution compared to MR, allowing for visualization of finer structures. Third, Spectral CTL eliminates patient contraindications that restrict DCMRL use, reduce scan time, and simplify workflows by permitting needle insertion in the CT environment and reducing the risk of needle dislodgement. However, spectral CTL introduces ionizing radiation absent with DCMRL. In order to ensure clinical implementations, low-dose static and dynamic protocols are a major prerequisite and have been optimized for other body regions (23–27). Moreover, advancements in CT technologies, such as the recent introduction of photon-counting CT, have further demonstrated potential dose reduction that can be exploited for spectral CTL (28–32). Currently, spectral CT technologies are primarily available at large academic and tertiary medical centers. However, continued clinical adoption of spectral CT may expand patient access to quantitative iodine mapping in the future. In addition, advanced reconstruction algorithms, such as deep learning reconstruction for both ultra-high resolution images and spectral images, have demonstrated significant noise reduction (33) and can help decrease dose needs for spectral CTL.

This study has limitations. First, the study only utilized two healthy animals to demonstrate feasibility and benefits of spectral CTL. Future studies will require additional animals to not only determine reproducibility but also evaluate the feasibility of detecting lymphatic pathological abnormalities prior to human studies. Second, animals were scanned with high dose to ensure good image quality for an initial assessment of spectral CTL. For future clinical translation, protocols must be designed to optimize dose for evaluating a range of lymphatic disorders. Existing CT technologies such as bolus tracking may be leveraged to standardize scan timing across a range of patients. Last, this study implemented spectral CTL with one type of spectral CT technology. Other spectral CT technologies, including photon-counting CT, also have iodine maps available and exhibited similar quantification capabilities critical for the benefits of spectral CTL (15–17,34,35).

Spectral CTL enhances existing lymphatic CT imaging with iodine maps that not only isolate contrast-enhanced lymphatic structures but also incorporate quantitative iodine density values for physiological characterization. The addition of quantitative metrics in non-invasive lymphatic imaging has the potential to augment current subjective assessment of central lymphatic structures to refine disease characterization and thus management.

## Data Availability

All data present in this study are available upon reasonable request to the authors.

## Acknowledgement

We acknowledge support through the National Institutes of Health (R01EB030494, R01EB035908) and Philips Healthcare.

## Notes

### Competing Interest Statement

Peter B. Noël has a research agreement with Philips Healthcare. Maxim Itkin is a stockholder of SureAx Medical and has educational support with Guerbet Inc.

